# Exploring Disparities and Novel Insights into Metabo-Nutritional Comorbidities among COVID-19 Patients in Mexico

**DOI:** 10.1101/2023.07.31.23293471

**Authors:** Ariel López-Cuevas, Mireya Martínez-García, Enrique Hernández-Lemus, Guillermo de Anda-Jáuregui

## Abstract

During the previous years, particularly at the beginning of the COVID-19 pandemic, the potential role of metabo-nutritional comorbidities in the severity and lethality of SARS-CoV2 infection has been widely discussed, often describing ambiguous outcomes. Here we investigate the prevalence of metabo-nutritional comorbidities among COVID-19 patients in Mexico. Using a retrospective observational study design, data was collected from official databases of COVID-19 patients admitted to public and private hospitals in Mexico City. Our study found a discordant prevalence of metabo-nutritional comorbidities among COVID-19 patients, particularly obesity, hypertension, and diabetes. Discordance consists in geographic location-dependent over and under-representation phenomena, that is the prevalence of such comorbidities in COVID-19 patients was significantly over or under the reported value for the general population in each location. These findings highlight the importance of screening for metabo-nutritional comorbidities in COVID-19 patients and suggest the need for tailored interventions for this population. The study also provides insights into the complex relationships between COVID-19 and metabo-nutritional comorbidities, which may inform future research and clinical practice.

## 1 Introduction

COVID-19, also known as the 2019 coronavirus disease, is a respiratory illness caused by the SARS-CoV-2 virus. It can cause a range of symptoms, from mild to severe, and can lead to serious complications in some people, particularly those who have underlying health conditions [33, 27, 19]. Metabolic conditions, such as obesity, diabetes, and hypertension (high blood pressure), are among the comorbidities that have been associated with worse outcomes in individuals with COVID-19 [4, 15]. A brief explanation of these main conditions and their potential association with COVID-19 outcomes follows:

### Obesity

Obesity is a condition characterized by an excess of body fat. It is often defined as having a body mass index (BMI) of 30 or higher. Obesity has been identified as a risk factor for severe COVID-19 and has been associated with an increased risk of hospitalization, intensive care unit (ICU) admission, and death. The mechanisms by which obesity may increase the risk of severe COVID-19 are not fully understood, but it is thought that obesity may cause inflammation and changes in immune function that make individuals more vulnerable to respiratory infections [25, 13].

### Diabetes

Diabetes is a chronic condition in which the body has difficulty regulating blood sugar levels. There are two main types of diabetes: type 1, which is an autoimmune disorder, and type 2, which is linked to lifestyle factors such as diet and lack of physical activity. People with diabetes, particularly those with uncontrolled blood sugar levels, are at higher risk for severe COVID-19 and have been found to have a higher rate of hospitalization and death compared to individuals without diabetes. It is thought that diabetes may impair the body’s ability to fight infections, leading to a greater risk of complications from COVID-19 [7, 6, 22, 14].

### Hypertension

Hypertension, or high blood pressure, is a common condition characterized by consistently high blood pressure readings. It can increase the risk of heart disease, stroke, and kidney disease. People with hypertension have been found to have a higher risk of severe COVID-19 and a higher rate of hospitalization and death compared to individuals with normal blood pressure. It is thought that hypertension may damage the blood vessels and increase the risk of serious complications from COVID-19 [28, 29, 20].

It is important to note that these metabolic conditions are just some of the comorbidities that have been associated with worse outcomes in individuals with COVID-19. Other comorbidities that have been linked to an increased risk of severe COVID-19 include cardiovascular disease, chronic lung disease, and immune system disorders. It is also worth mentioning that while these conditions may increase the risk of severe COVID-19, it is not necessarily the case that all individuals with these conditions will experience severe COVID-19. The overall risk of severe COVID-19 and the likelihood of complications depend on a variety of factors, including the individual’s age, overall health, and underlying medical conditions [11, 9, 2].

These conditions are also prevalent in Mexico, and research has shown that they may increase the risk of severe COVID-19 and worse outcomes in Mexican populations.

Obesity is a major health concern in Mexico, with more than 70% of adults classified as overweight or obese. Obesity has been identified as a risk factor for severe COVID-19 and has been associated with an increased risk of hospitalization, intensive care unit (ICU) admission, and death in Mexican populations. In a study of COVID-19 patients in Mexico, obesity was found to be a significant predictor of severe COVID-19, with obese patients having a 3.7 times higher risk of severe disease compared to non-obese patients. Diabetes is also a major health concern in Mexico, with nearly 14% of the adult population affected. People with diabetes, particularly those with uncontrolled blood sugar levels, are at higher risk for severe COVID-19 and have been found to have a higher rate of hospitalization and death compared to individuals without diabetes in Mexico. In a study of COVID-19 patients in Mexico City, diabetes was found to be associated with a higher risk of severe COVID-19 and death. People with hypertension have been found to have a higher risk of severe COVID-19 and a higher rate of hospitalization and death compared to individuals with normal blood pressure in Mexico. In a study of COVID-19 patients in Mexico City, hypertension was found to be associated with a higher risk of severe COVID-19 and death [23].

It is worth mentioning that while these conditions may increase the risk of severe COVID-19 in Mexico, it is not necessarily the case that all individuals with these conditions will experience severe COVID-19. The overall risk of severe COVID-19 and the likelihood of complications depend on a variety of factors, including the individual’s age, overall health, and underlying medical conditions. In order to have a deeper understanding of how these comorbidities have affected the populations’ risks in the context of some environmental constraints, we will perform further statistical analysis in geographically adjacent clusters.

## 2 Methods

### Data source and sample selection

We obtained clinical and demographic data for our study from the Sistema Nacional de Vigilancia Epidemiológica (SINAVE, https://www.gob.mx/salud/documentos/datos-abiertos-152127), which is a COVID-19 database managed by the Mexican federal health authorities. Our study included all positive cases (identified by either PCR or antigen testing) reported in SINAVE up to epidemiological week 2022-39. We included cases from all municipalities in Mexico.

### Data collection and classification

We extracted information on the reported status of metabonutritional comorbidities, including obesity, diabetes, hypertension, and dyslipidemia, from the SINAVE database. We classified cases into three outcomes: ambulatory, hospitalized, or deceased, based on the information reported in the database. We also subdivided the cases into waves following the wave definition proposed by Sifuentes-Osornio et al. (2023).

### Variables and Analysis

We considered the reported status for metabonutritional comorbidities for each case in SINAVE, and stratified them into those with obesity, diabetes, hypertension. We then calculated the fraction of these comorbidities among COVID-19 cases for each municipality, considering the aforementioned subdivision in ambulatory, hospitalized or deceased groups.

### Metabonutritional comorbidity prevalence

We obtained data on obesity, diabetes, and hypertension at the municipal level from the Estimación para Áreas Pequeñas (EAP), made by the National Institute of Public Health (INSP) and the National Institute of Statistics and Geography (INEGI) using the Encuesta Nacional de Salud y Nutrición (ENSANUT) 2018. We used a public version of this dataset made available in https://github.com/rojoneon/ensanut_mun.

### Over and underrepresentation analysis

Over or under representation was defined as having a fraction of COVID-19 cases with the comorbidity with a difference of 5% or more with regard to the reported prevalence. For example, if the prevalence of obesity in a certain municipality was 30%, and the fraction of COVID-19 cases with obesity was 40%, then that municipality was considered overrepresented. Conversely, if the prevalence of obesity was 30%, and the fraction of COVID-19 cases with obesity was 20%, then that municipality was considered underrepresented.

## 3 Results and discussion

In this work we explored the relationship between the prevalence of metabonutritional comorbidities and the incidence and outcomes of COVID-19 in each municipality of Mexico. First, we looked at the spatial distribution of these comorbidities, and compared them to the all-time incidence and mortality of COVID-19; showing differences in their spatial distribution. We then compared the prevalence of this comorbidities in the general population with their frequency in COVID-19 patients; showing divergences where some comorbidities are over-represented while others are under-represented. These patterns of over and under representation change in time. Finally, we look into the role of testing volume as a possible explanation for these divergences.

### 3.1 Geographical distribution of COVID-19 and its nutrimetabolic comorbidities

The maps shown in figures 1 and 2 showcase the spatial distribution of incidence and mortality of COVID-19 according to the public data released by SINAVE. On the other hand, the the maps in figures 3 through 5 show the spatial distribution of the prevalence of metabo-nutritional comorbidities, according to the ENSANUT data. This maps show that the patterns of spatial distribution for COVID-19 (both incidence and death rates) and the prevalence of metabo-nutritional comorbidities are different.

**Figure 1:**
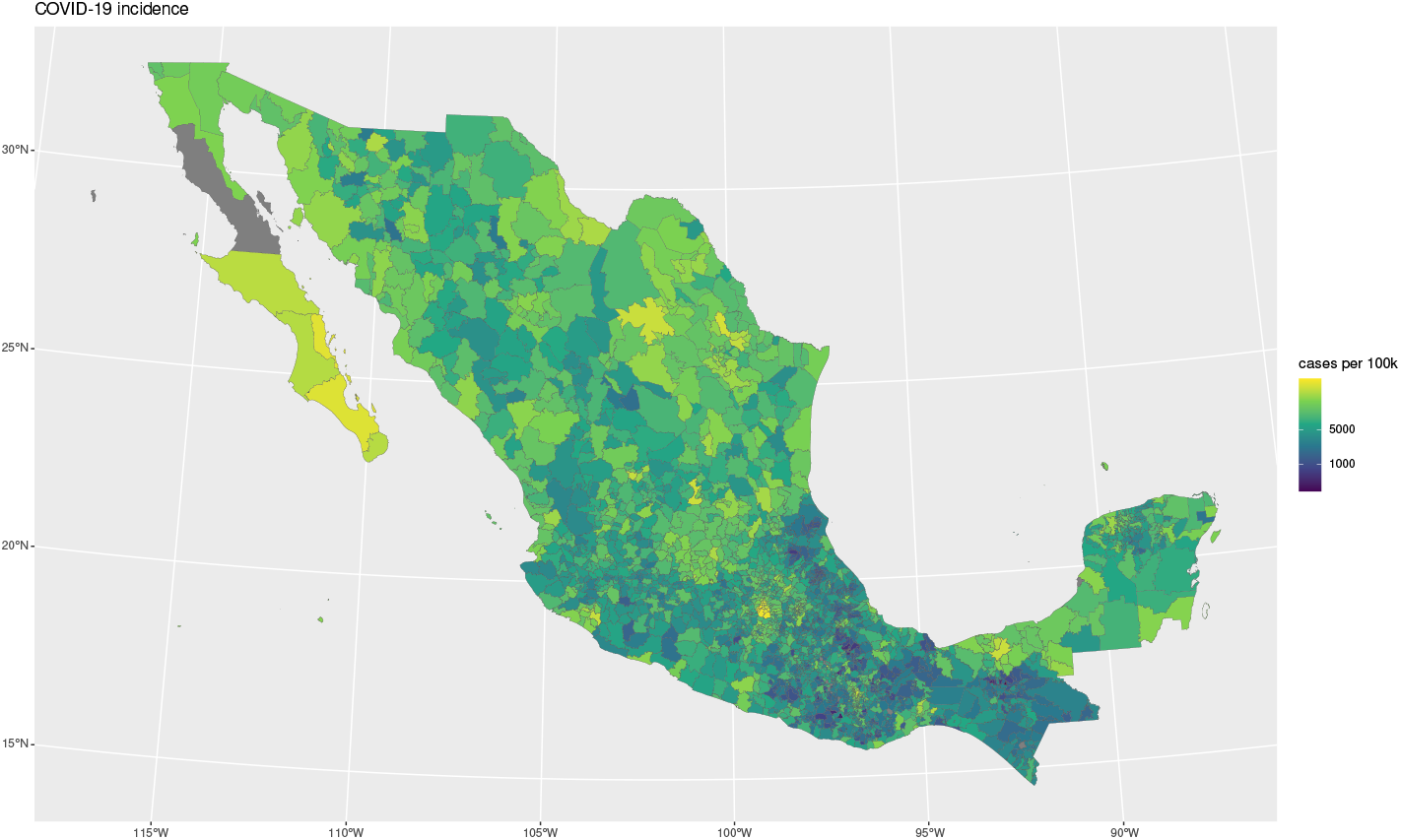
COVID-19 incidence

**Figure 2:**
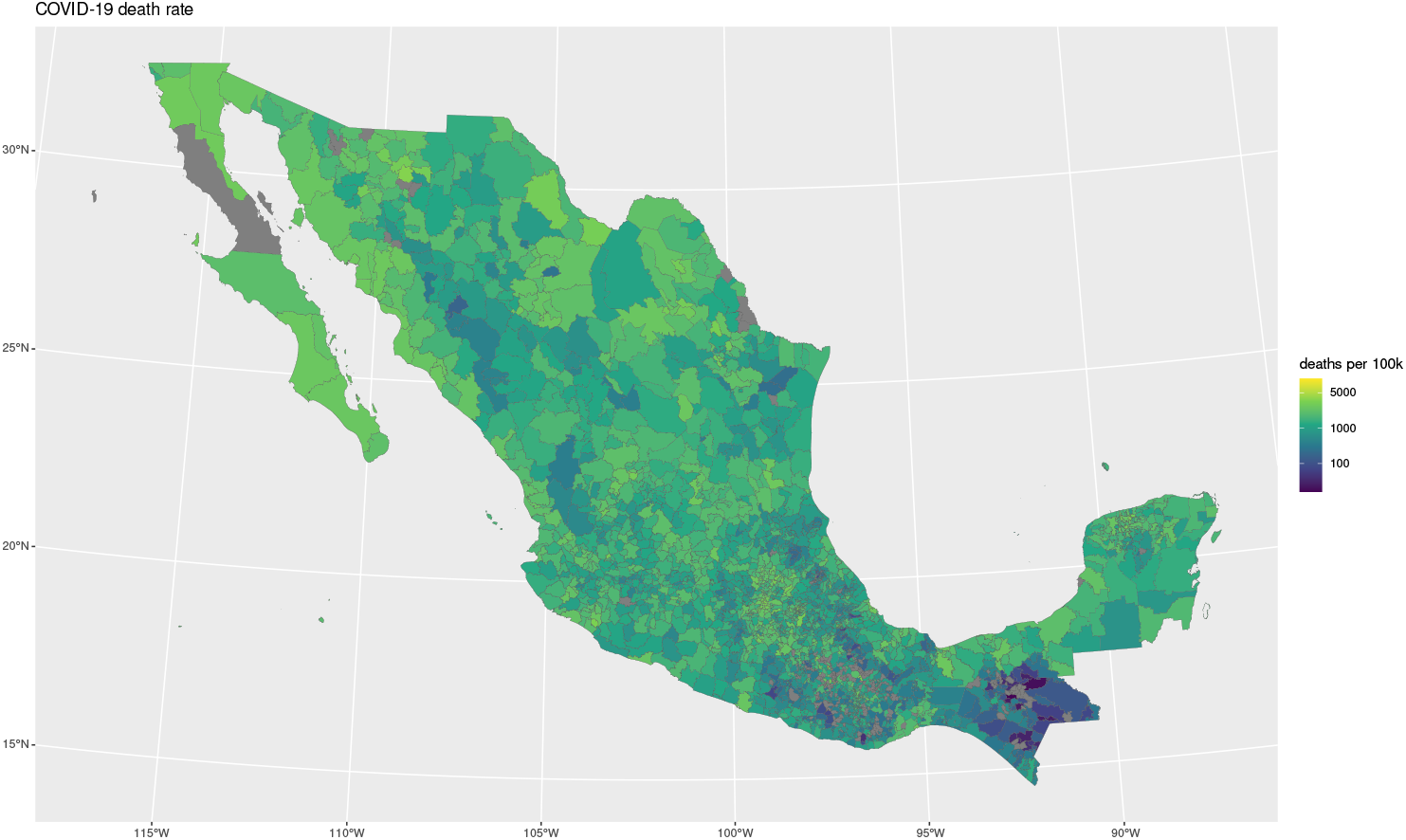
COVID-19 death rate

**Figure 3:**
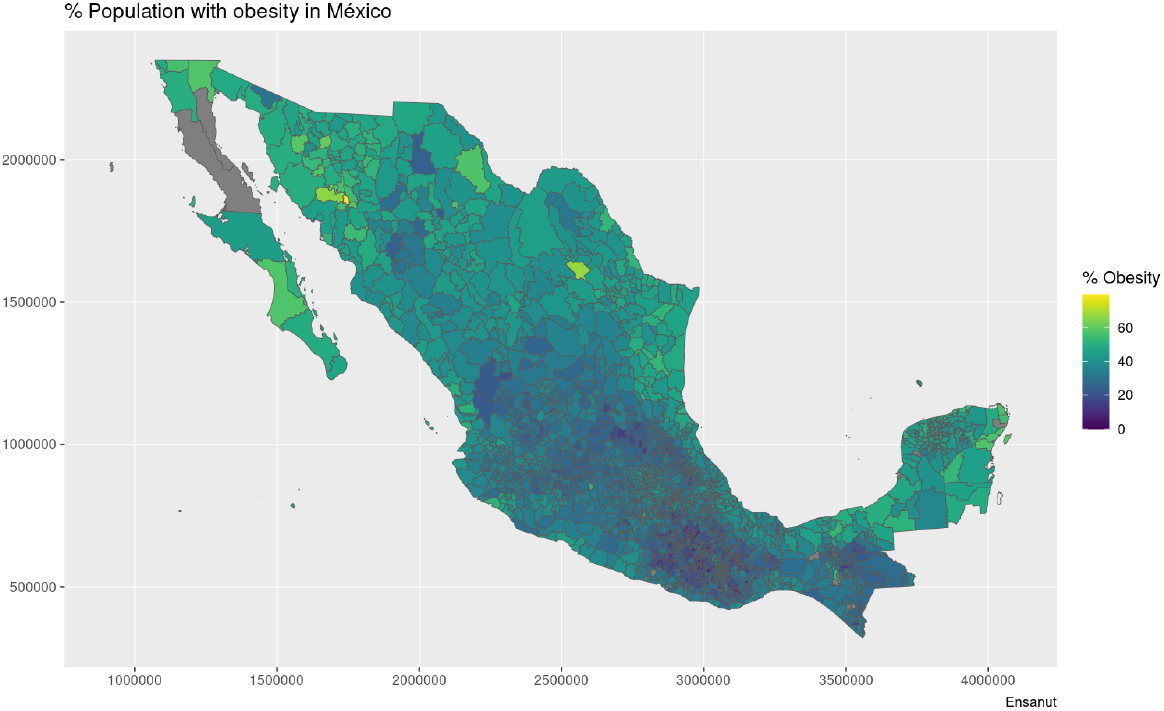
Municipality-wise distribution of Obesity prevalence in Mexico

**Figure 4:**
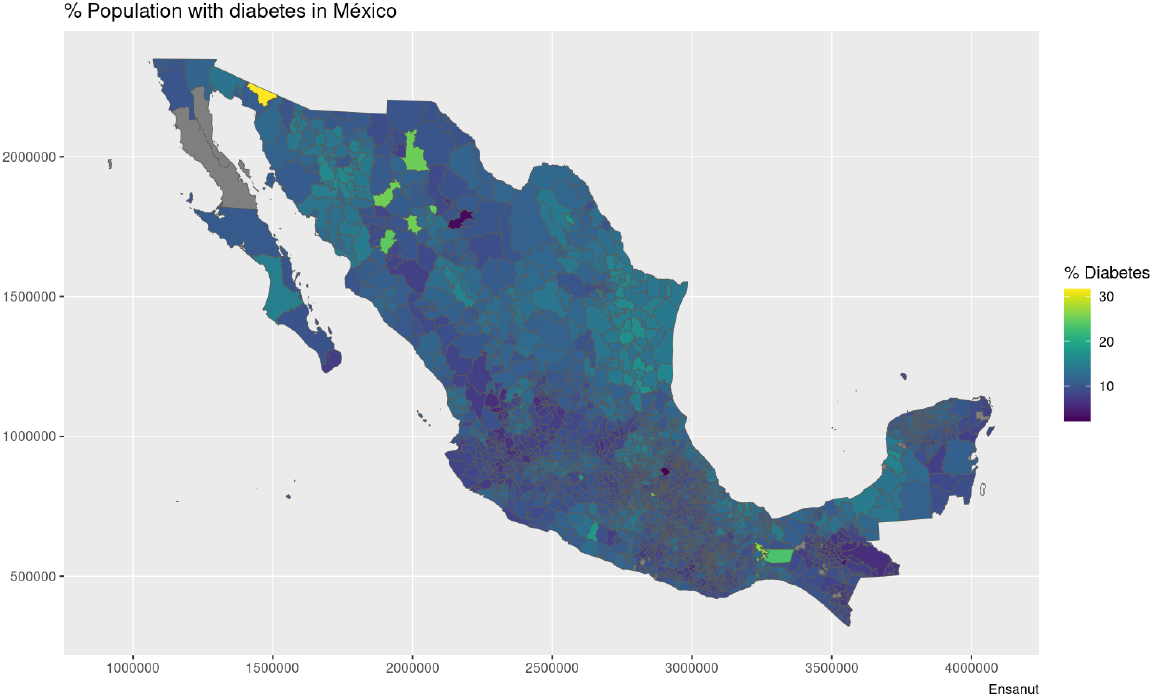
Municipality-wise distribution of Diabetes prevalence in Mexico

**Figure 5:**
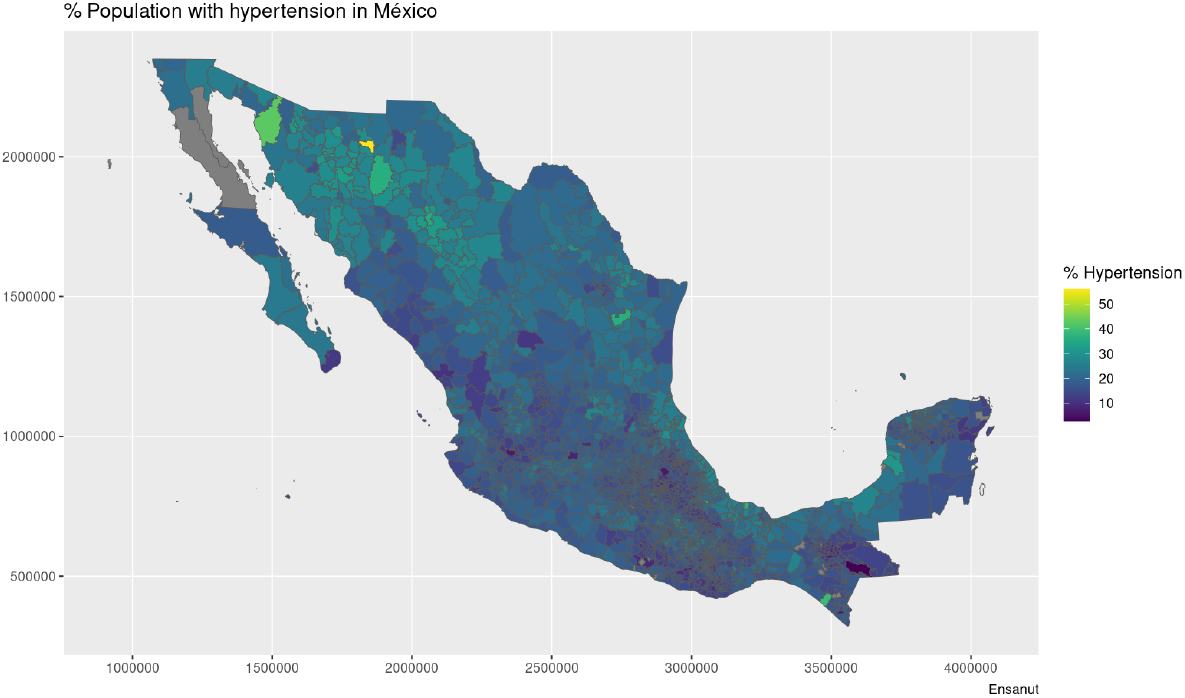
Municipality-wise distribution of Hypertension prevalence in Mexico

**Figure 6:**
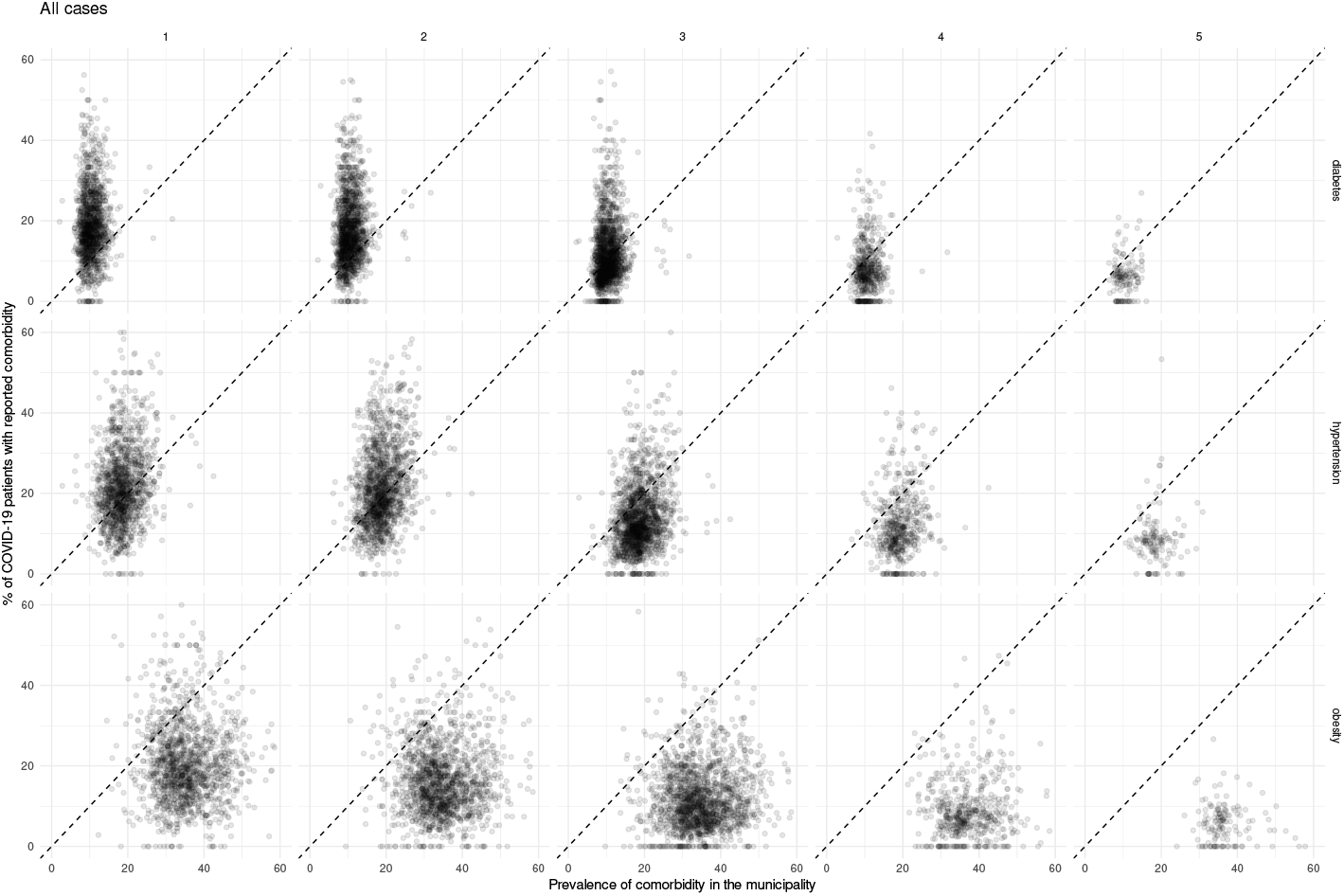
Scatterplots showing mexican municipalities according to comorbidity prevalence and fraction of all COVID-19 patients with the given comorbidity for each epidemic wave.

**Figure 7:**
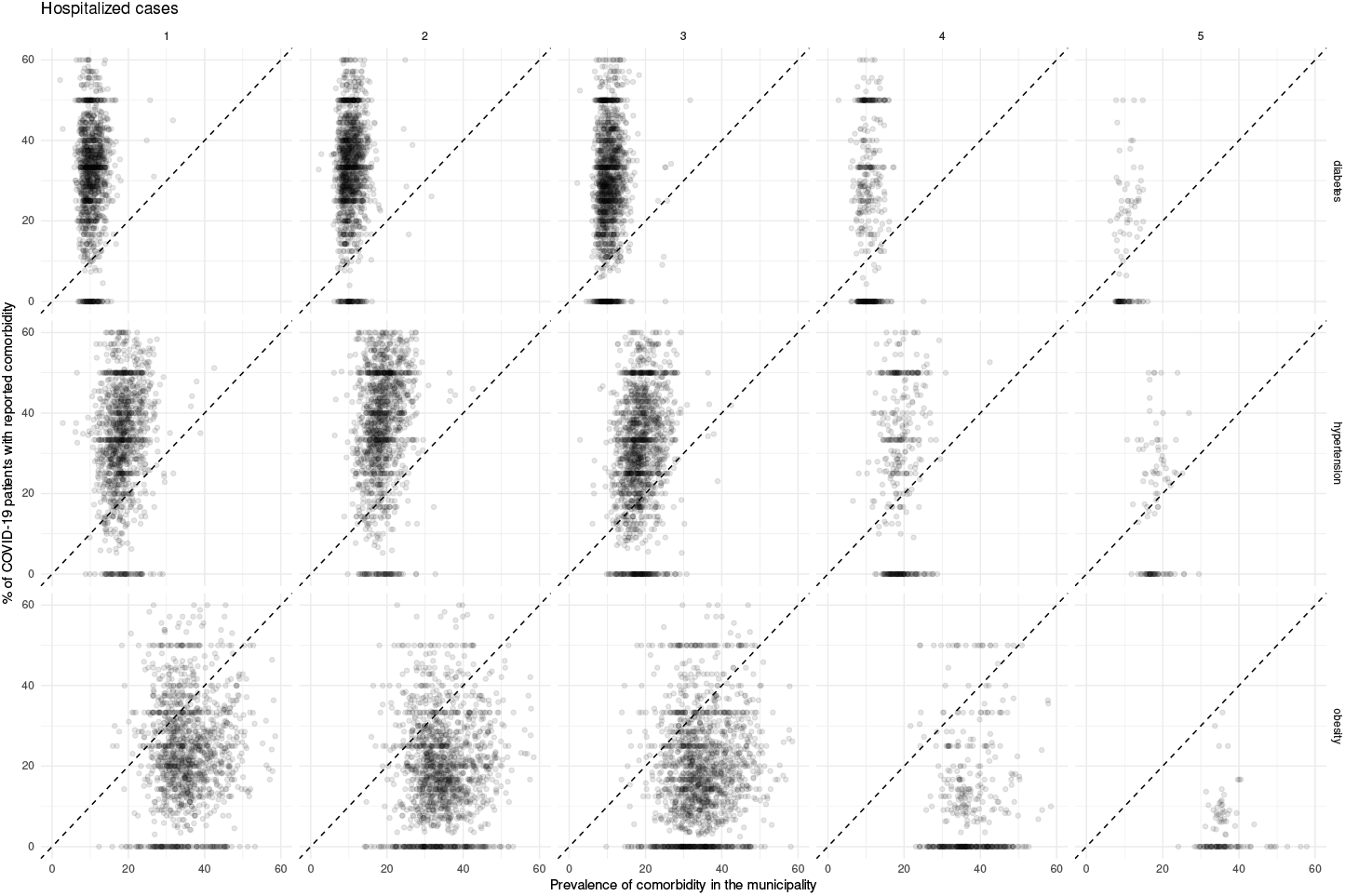
Scatterplots showing mexican municipalities according to comorbidity prevalence and fraction of COVID-19 patients requiring hospitalization with the given comorbidity for each epidemic wave.

**Figure 8:**
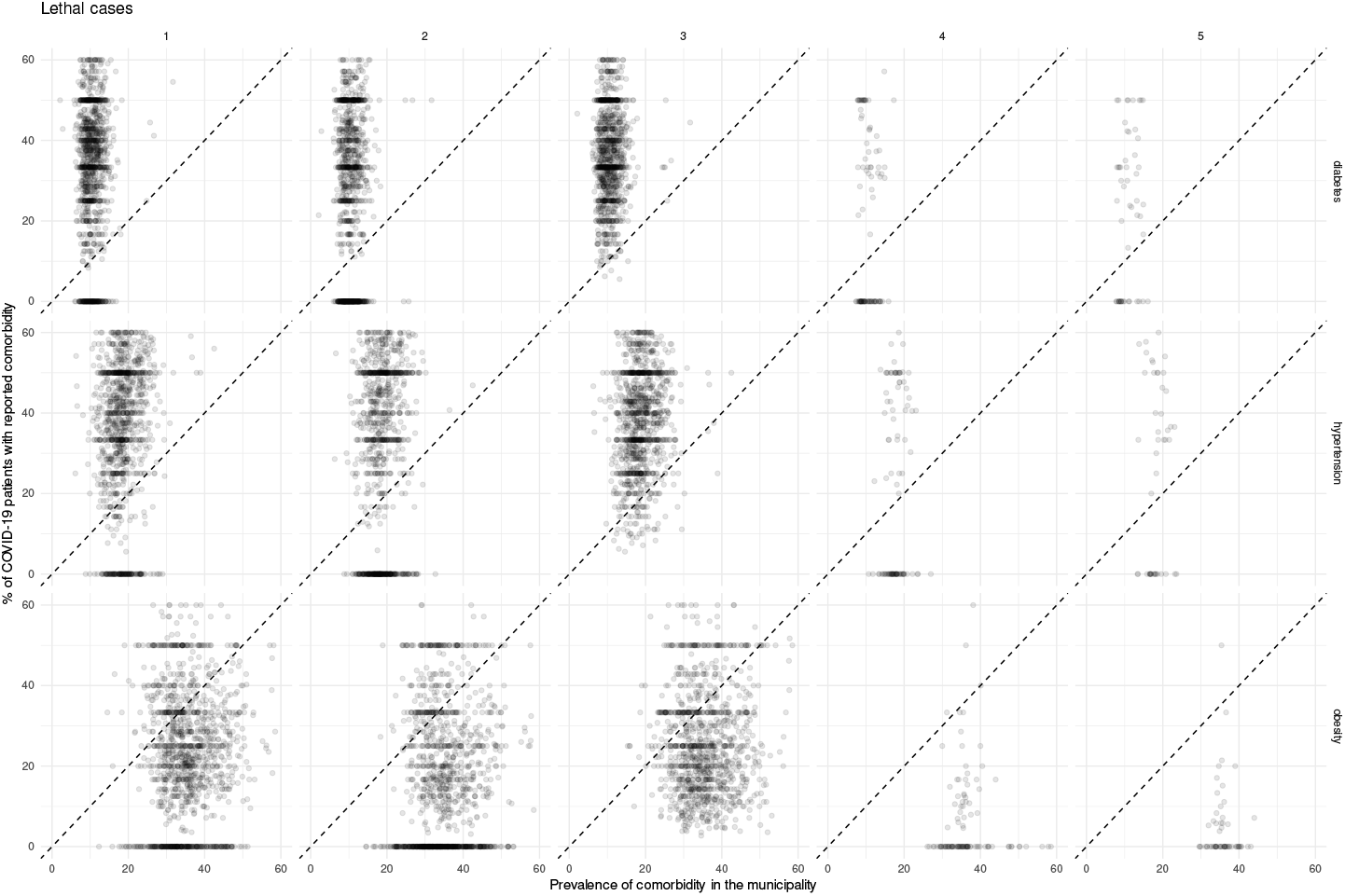
Scatterplots showing mexican municipalities according to comorbidity prevalence and fraction of lethal COVID-19 cases with the given comorbidity for each epidemic wave.

**Figure 9:**
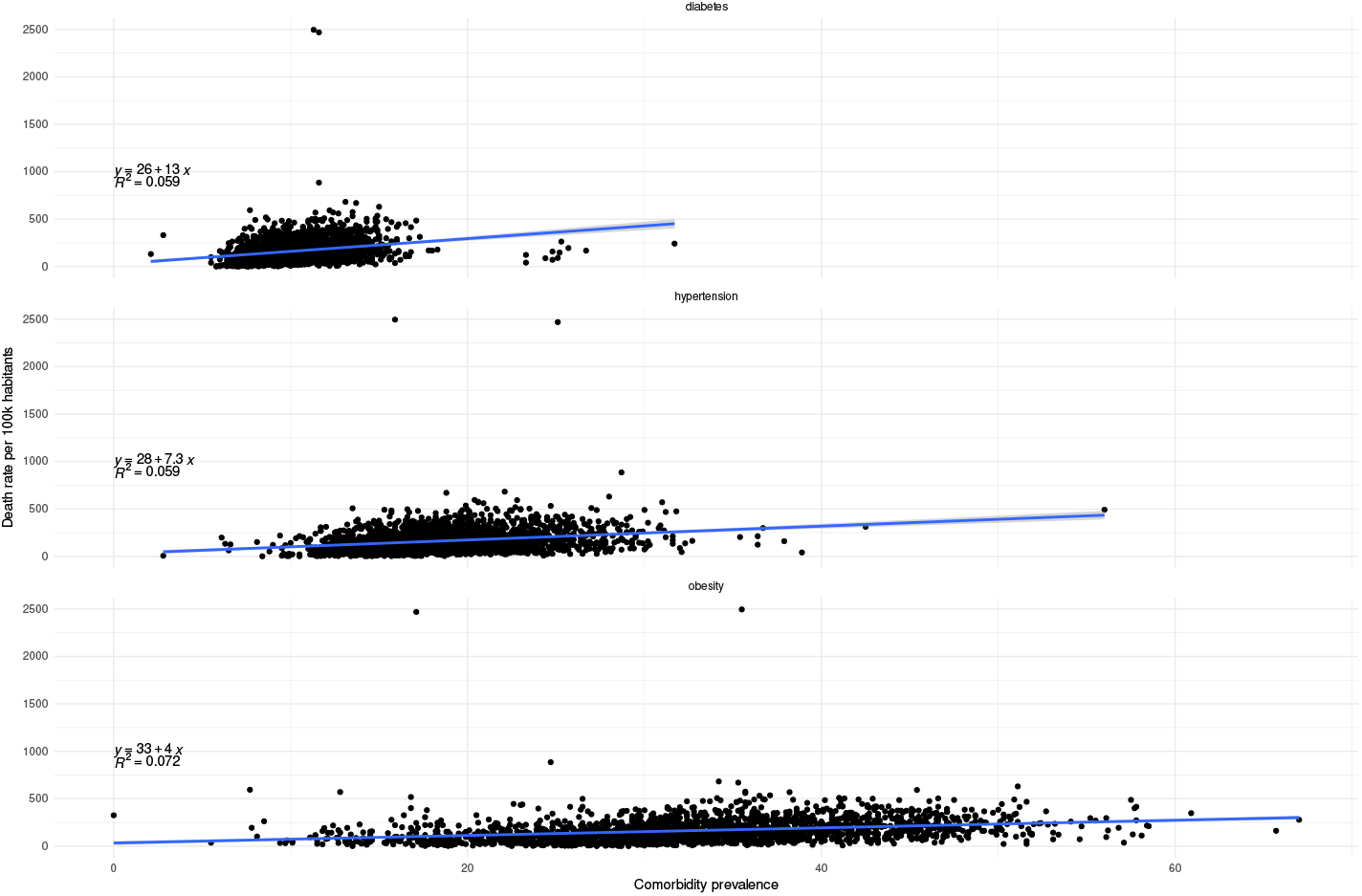
Correlation between comorbidity prevalence and mortality at the municipality level.

**Figure 10:**
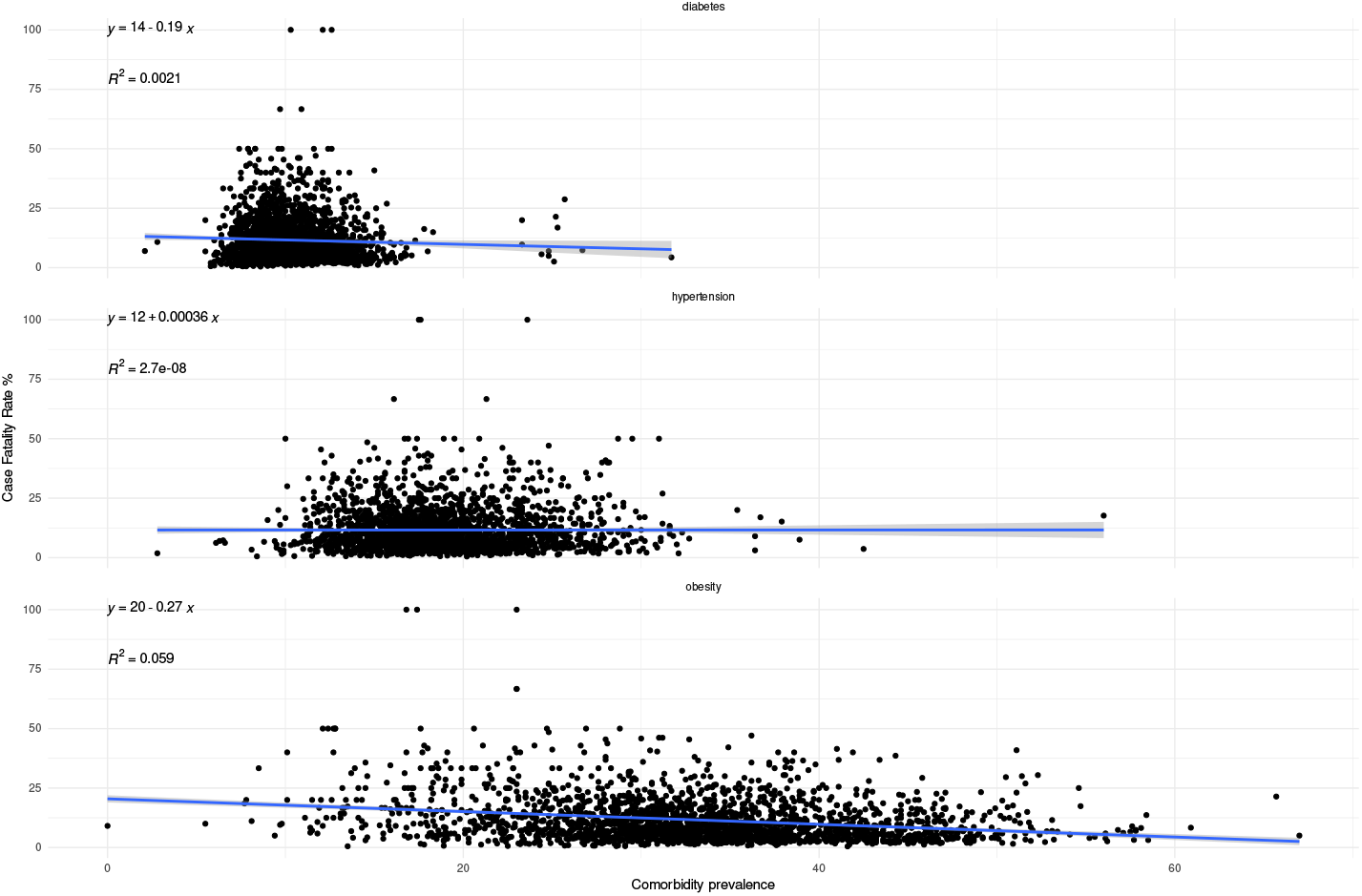
Correlation between comorbidity prevalence and lethality at the municipality level.

**Figure 11:**
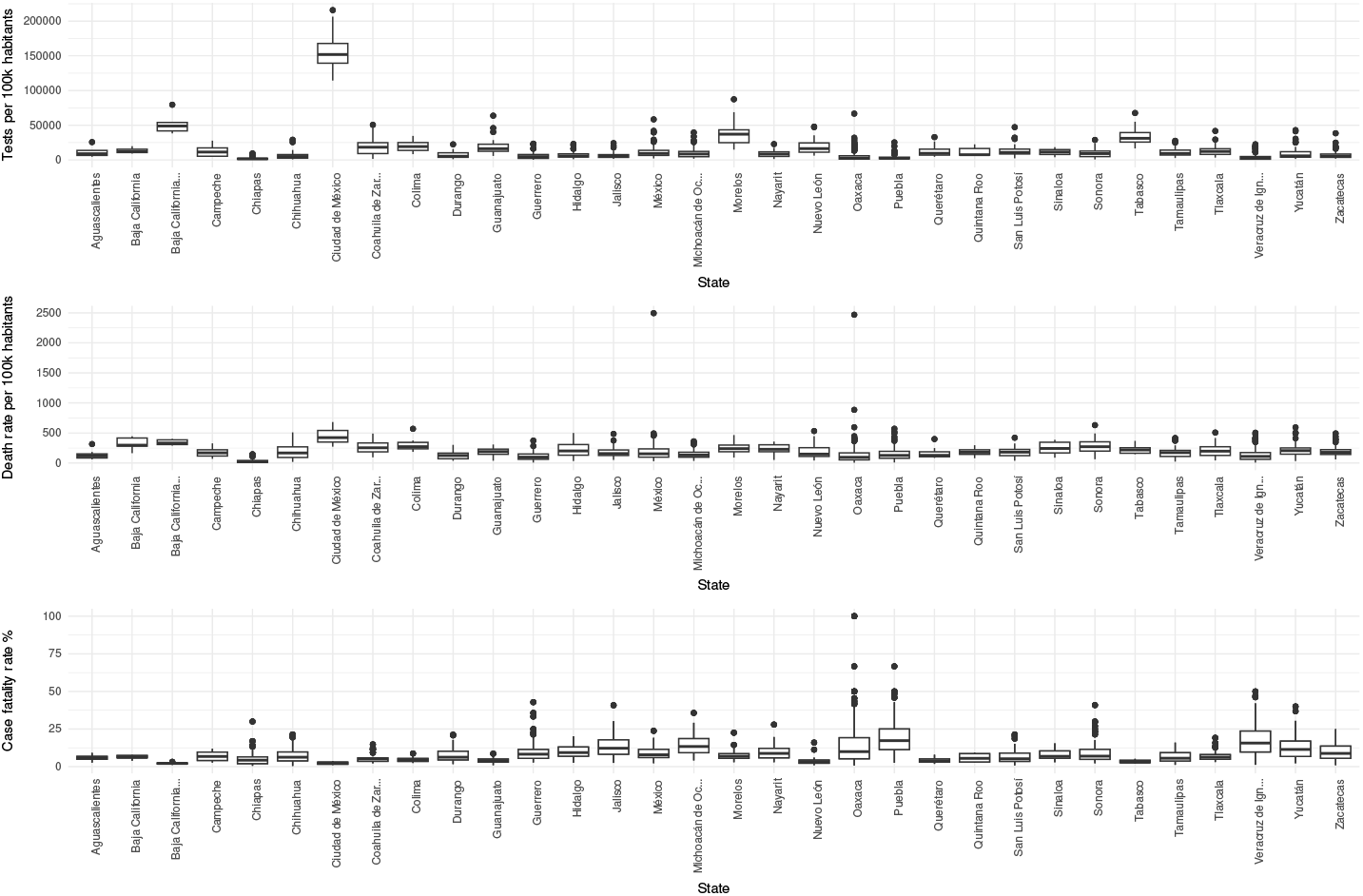
Boxplots showing the distributions of test rates, mortality and lethality per state. It highlights that states with higher testing rates generally detect more lethal cases, but register a lower lethality.

**Figure 12:**
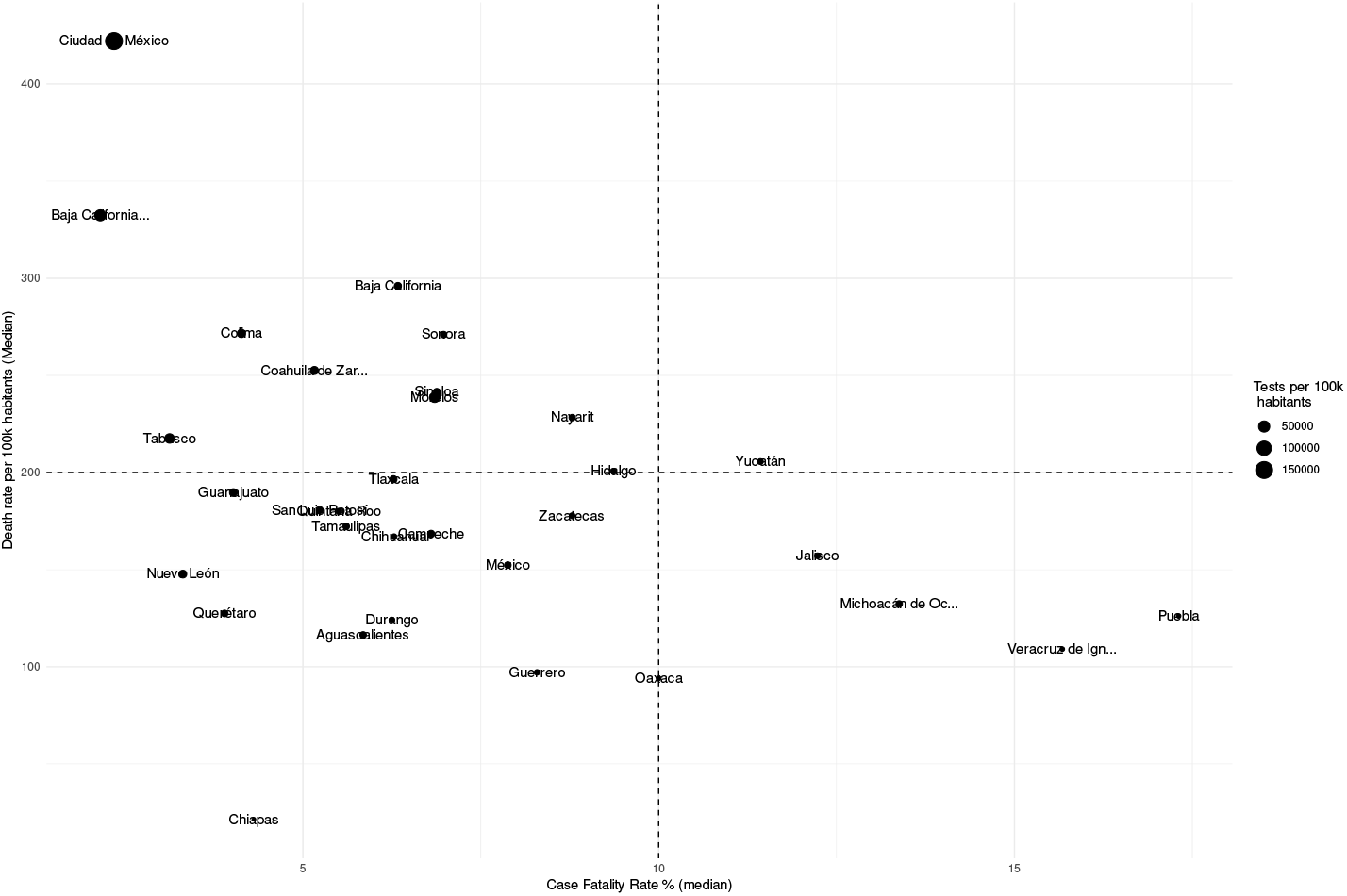
Scatterplot showing the relationship between mortality, lethality and test rates per state.

**Figure 13:**
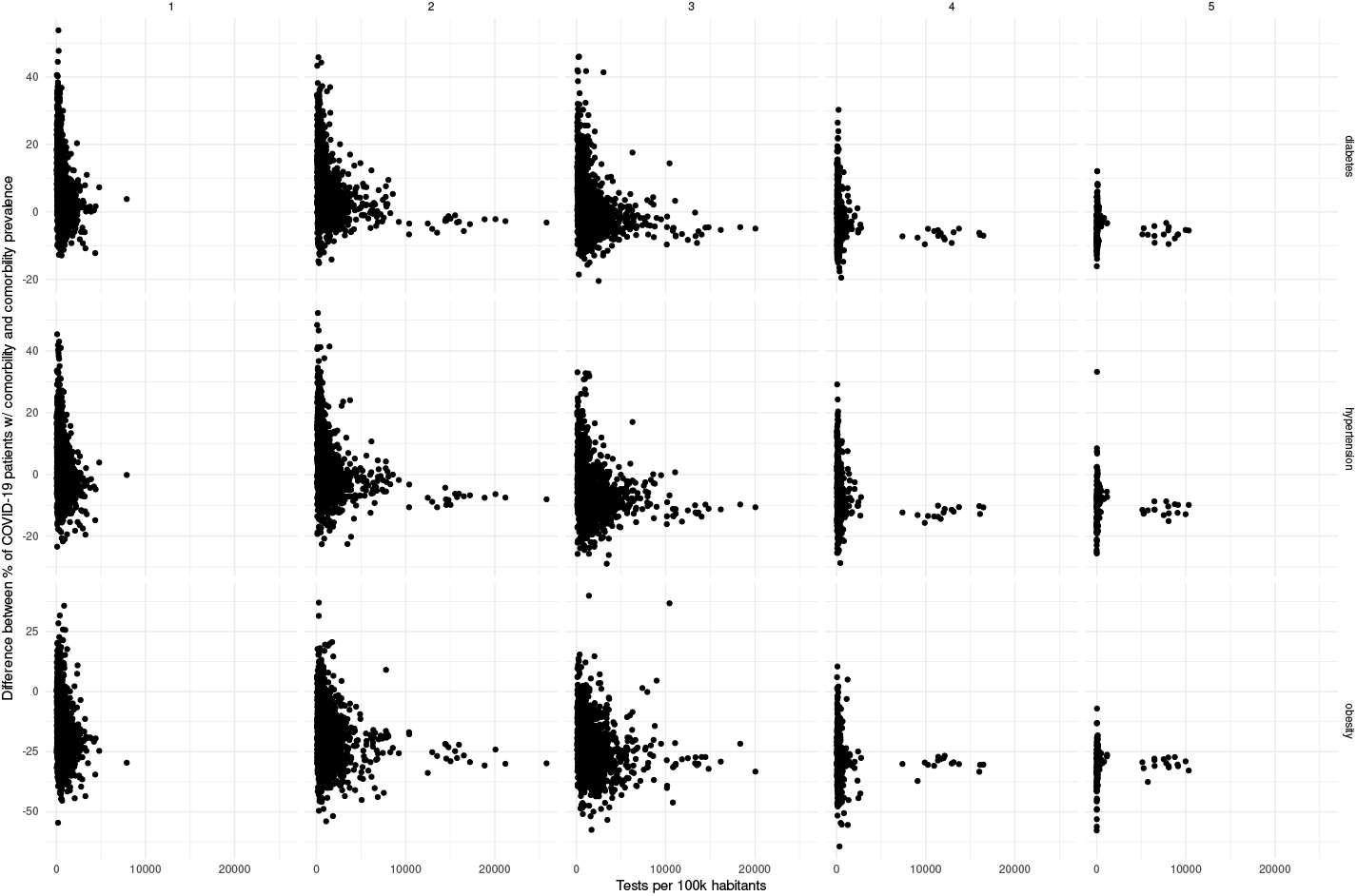
Scatterplot of testing volume vs divergence between comorbidity representation in COVID-19 patients and prevalence.

At first glance, some north and south divides may be observed for diabetes and hypertension — with higher prevalences in the northern states. Meanwhile, in the case of COVID-19, both incidence and death rates exhibit less definitive patterns; the exception being the states of Veracruz, Oaxaca and Chiapas, which exhibit lower incidences and death rates. Notice that in the case of the COVID-19 maps, the color scale is in a pseudolog scale. For a better viewing experience, we are providing these maps in vector graphics in Supplementary Material 1.

Metabolic and cardiovascular comorbidities, such as obesity, diabetes, and hypertension, are strongly influenced by social determinants of health (SDHs) that encompass various social, economic, and environmental factors affecting an individual’s well-being beyond genetic predispositions [24, 26, 31, 16, 32]. In Mexico, the significant problem of social inequity exacerbates the impact of SDHs on the population’s health dynamics. Marginalized and disadvantaged populations bear a disproportionate burden of metabolic comorbidities due to limited access to healthy foods, healthcare, and other vital resources [12].

Considering that social determinants have also been associated to COVID-19 incidence and negative outcomes [1, 18], the question remained on whether the frequency of these comorbidities in COVID-19 cases reflected the prevalence of the diseases in the general population.

### 3.2 Representation of comorbid populations within the COVID-19 patient populations

COVID-19 is an infectious disease that is transmitted primarily through contact between people, such that virions in respiratory particles can be transferred from an infected patient to a susceptible individual [21]. Considering a traditional epidemiological scenario in which a given population exhibits homogeneous mixing [30], a naive assumption would be that the infected population would be a random sample of the general population. In that case, it would be expected that the fraction of cases that exhibit a given condition, such as a comorbidity, would be the same than in the larger population:

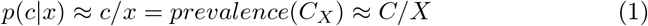

That is, the probability of a case with the comorbidity (*c*) in the infected population *x* (which, if the epidemiological surveillance system is adequate, should approach *c*/*x*) should be the same as the prevalence of the comorbidity *C* in the population *X* (*C*_*X*_), which properly measured should yield the number of comorbidity cases over the population size.

For this to be true, however, the risk of infection should be independent from the presence of the comorbidity (that is, the comorbidity should not confer a mechanistic susceptibility that increases the capacity of the virus to infect an individual with the comorbidity). With this in mind, a deviation from this equality would mean (at least) one of the following:

- The epidemiological surveillance system is inadequate.
- The prevalence estimation is inadequate.
- *p*(*c*|*x*) ≠ *p*(*c*|*x*); that is, there is a mechanism that makes the risk of infection for the comorbid population different than that for the non-comorbid population.

Since a fraction of the infected population will have negative outcomes such as hospitalization and death, a similar mathematical argument can be established. In this case, however, it has been proven that certain metabo-nutritional comorbidities do increase the risk of hospitalization and death in COVID-19 patients [23]; therefore, over-representation would be expected.

With this in mind, we explored the representation of comorbidities in each municipality for COVID-19. We present this analysis broken down by waves (defined in terms of changes in hospitalization demand as defined in [23]), as there are well documented differences in social conditions (ie. lockdowns), resources (such as testing), and the implementation of vaccination makes between these time frames.

In what follows we present and discuss these divergences using scatterplot visualizations, in which a diagonal dotted line provides a visual guide for over-representation (dots over the line) and under-representation (dots under the line). Additionally, we provide the data used for these plots (along with numerical over-representation analysis) in Supplementary File 1.

For all COVID-19 cases, we observe over-representation of both diabetes and hypertension in the majority of municipalities in the early waves of the pandemic; such over-representation decreases in later waves. On the other hand, throughout the pandemic there was a sustained under-representation of obesity in COVID-19 cases; to the point that in the fifth wave there was no municipality with an over-representation of obesity.

Meanwhile, for both hospitalization and deaths, the over-representation of diabetes and hypertension is sustained throughout the pandemic. Whereas obesity does exhibit a shift, where more municipalities exhibit under-representation as the pandemic advances.

#### 3.2.1 Relationship between comorbidities and lethality, death rate, and testing

The role of comorbidities in complications among individuals in Mexico has been extensively studied [10, 5] in the scientific literature. However, inaccurate confusion between this risk and the population risk due to prevalence have permeated in the public discussion, without noting the divergence discussed in this manuscript. With this in mind, we now turn our attention to exploring the relationship between the population prevalence of comorbidities and the rates of mortality and lethality. For the purposes of self-containment, we will assess this measures based on deaths recorded in the SISVER public database, which shows an undercounting compared to excess mortality estimations [3].

As we may see, the correlation between prevalence and both mortality and lethality is rather week. However, it is interesting to observe that the correlation trend is positive for mortality for all comorbidities, but negative for lethality in the case of diabetes and obesity.

An explanation may be found in testing patterns. Briefly, if the testing base is large, then a larger set of non-lethal cases may be found, which reduces the lethality; however, as more cases are detected, more deaths are counted, leading to a higher mortality. In Supplementary Material 1 we present the relationship between lethality, death rate and testing, using boxplots for each measure (grouping the municipalities by state), as well as a scatterlot combining the three measures. We may see that in terms of all three measures Mexico City is the state with the higher testing rates, which increased its mortality (since more lethal cases were confirmed) but greatly reduced the case fatality rate.

Interestingly, this circles back to the representation: in the last figure, we may observe that the municipalities that performed the largest number of tests consistently show underrepresentation of the comorbidites. However, large testing is not necessary for underrepresentation; that is, there may be other mechanisms behind divergences in representation.

## 4 Limitations, conclusions, and future work

This work attempts to provide information regarding the discrepancies between the frequency of comorbidities between COVID-19 cases and the general population. Certain limitations should be acknowledged. One is that the estimation of prevalence at the municipal level was generated using ENSANUT 2018 data, which may be outdated. When a new estimation is available, this data should be reanalyzed.

Another limitation is that, as previously mentioned, the death counts in the SISVER database are underestimated. However, for the analysis presented in this work we must rely on this data, as other sources lack information on comorbidities, which is at the core of this work.

Finally, we shall emphasize that, while we worked around the traditional assumption of homogeneous mixture, infectious agents like SARS-CoV-2 spread through complex networks [17]. Reconstructing such networks for mexican populations is an ongoing endeavour [8]. Considering such heterogeneous patterns may capture some of the differences in representation, particularly in terms of infection risk. This may be important for modelling purposes for this and future pandemic, as it may point to differentiated decision-making and risk-assessment in risk populations. However, this is beyond the scope of the current manuscript.

In conclusion, here we show that the metabonutritional comorobidities that have been identified as important for risk of unfavourable outcomes in COVID-19 patients, showed frequencies not expected by their general population prevalence. We show that this prevalence is not sufficient to explain trends in lethality and mortality. And we propose that testing patterns may be one of the factors that explain these divergences.

## Data Availability

All data produced in the present work are contained in the manuscript

## Author Contributions

ALC: Data analysis, code, visualization, discussion, writing. MMG: Data analysis supervision, discussion. EHL: Statistical analysis, discussion, writing, design. GDJ: Data analyisis, code, discussion, writing, design.

## Acknowledgments

The authors would like to thank Máximo Ernesto Jaramillo-Molina for making the metabo-nutritional prevalence data at the municipal level publicly available in an open format.

ALC is a student at the Autonomous University of Mexico City (UACM).

